# Management and outcomes of fractures over cranial venous sinuses: A scoping review protocol

**DOI:** 10.1101/2023.11.20.23298713

**Authors:** Berjo D. Takoutsing, Ubraine Wunde Njineck, Praise Senyuy Wah, Lorraine Sebopelo, Josiane Ngouanfo Tchoffo, Ella Shiynyuy Bah, Ignatius Esene

## Abstract

**Introduction:** Fractures over venous sinuses (FOVS) are associated with difficulties in diagnosis, and treatment resulting in a high level of morbidity, and mortality. Despite its importance, there is limited aggregate data to guide the management of these fractures ultimately inflicting a major challenge to neurosurgeons. This protocol describes the methodology of a scoping review that aims to synthesise contemporary evidence on the management and outcomes of FOVS.

**Methods and analysis:** The proposed study will be conducted in accordance with the Arksey and O’Malley’s framework for scoping reviews. The research question, eligibility criteria and search strategy were developed based on the Population, Intervention, Comparator, Outcome strategy. The following electronic bibliographic databases will be searched without restrictions on language, and date of publication: PubMed, WHO Global Index Medicus, African Journals Online, SCOPUS, Embase. All peer-reviewed studies of primary data reporting on the management and outcomes of FOVS will be included. The data extracted from included articles will be presented through descriptive statistics, pooled statistics, and a narrative description.

**Ethics and dissemination:** Because this study did not directly involve human individuals, ethical approval was not necessary. Dissemination strategies will include publication in a peer-reviewed journal, oral and poster presentations at local, regional, national and international conferences and promotion over social media.

**ARTICLE SUMMARY:** *Strengths and limitations of this study:*

- To our knowledge this is the first scoping review focusing on the management and outcomes of fractures over the venous sinuses
- This protocol will ensure transparency in the methodology of the proposed review, thus will reduce the likelihood of reviewing bias.
- The search strategy will be conducted in five electronic databases commonly used across both high- and low-income countries.
- There will be no restrictions on language, location, or publication date during the screening process.
- Unpublished studies will not be sought.
- There will be no formal assessment on the quality of the included studies.

## INTRODUCTION

Skull fractures following trauma continues to be a major public health problem disproportionately affecting low- and middle-income countries (LMICs).^1^ It is commonly associated with traumatic brain injury (TBI) occuring in 28-37% of affected patients.^1,2^ Several types of skull fractures exist, with depressed skull fractures and skull base fractures usually associated with injury to the underlying dural venous sinuses.^3^ Despite its importance, fractures over venous sinuses (FOVS) can be associated with a missed or delayed diagnosis, and difficult management.^4^ These results in a high morbidity, and mortality in the pre-operative, operative and perioperative periods.^5^

About 10% of skull fractures occur over the cranial venous sinuses, with the majority occurring in the context of direct trauma, followed by road traffic accidents especially in LMICs owing to safety measures not fully implemented.^6,7^ FOVS are associated with high in-patient, mixed clinical presentation and are prone to high mortality.^5,8^ The management can be conservative or surgical both of which have different indications and risks.^6^ Due to the limited guidelines or consensus on the management of these fractures coupled with the unavailability of imaging modalities such as CT scans, MRI in resource limited settings, neurosurgeons are thus faced with a therapeutic dilemma usually relying on good judgment and experience.^9^

There are ongoing investigations on novel therapeutic interventions to reduce mortality associated with FOVS.^8,10^ However, they may be inaccessible in resource-limited settings and emergency cases. There is therefore a need for a comprehensive overview of the available literature on the management and outcomes of these fractures to identify the various therapeutic modalities, assess the availability of various diagnostic methods, and evaluate patients’ outcomes. This would fill in key research gaps and priorities, and inform future interventions.

## STUDY AIMS

This protocol describes the methodology of a scoping review of the literature on the management and outcomes of cranial venous sinus fractures. **Box 1** describes the primary and secondary goals of the proposed review. The suggested review will provide information to health care practitioners, policymakers, stakeholders, and healthcare organisations involved in providing care to patients who have suffered venous sinus fractures. The knowledge gained will aid in the identification of key priorities and the establishment of larger research initiatives with a greater level of evidence.

### Box 1.

Primary and secondary aims of the scoping review

Primary aim:

1. Describe the therapeutic modalities for FOVS.

Secondary aim:

2. Assess the availability of diagnostic methods including neuroimaging (MRI, CT scan), and fundoscopy.
3. Evaluate patient’s clinical outcomes primarily defined as mortality and complication rates, Glasgow Outcome Score, Length of hospital stay, and clinical state on discharge.

## METHODS

The planned scoping review will be carried out in accordance with the five stages of Arksey and O’Malley’s framework.^11^ This review’s protocol is formatted in accordance with the Preferred Reporting Items for Systematic Reviews and Meta-Analyses (PRISMA) of Protocols.^12^

### Stage 1: identifying the research question

The Patient, Intervention, Comparison, Outcome (PICO) framework was utilised to determine the title, objectives, research questions, and eligibility criteria for the scoping review. Our key research question is: How are venous sinus fractures managed globally?

### Stage 2: identifying relevant studies

The authors defined the keywords that will be employed in the search strategy as well as the study eligibility criteria using the PICO approach.

### Databases and search strategy

BDT developed the search strategy to identify research papers on fractures over the venous sinus using multiple combinations of keywords, MeSH terms, and different electronic bibliographic databases. Following discussion with the review team, the final search strategy broadly consisted of variants of ‘Skull fracture’ and ‘Venous sinus’ (**Supplementary Table 1**). The literature search was conducted from inception until 29 October 2023 without restrictions on language or date of publication in the following databases: PubMed, WHO Global Index Medicus, African Journals Online, Scopus, and Embase. A reviewer will hand search additional resources such as Google scholar, and references from included studies to verify that all relevant material in the literature is included.

### Eligibility criteria

An eligibility criteria was developed following discussion with the reviewers to guide the screening phase. The inclusion criteria adhere to the PICO strategy and are listed below:

#### Population

All studies of patients of all ages with a diagnosis of venous sinus fracture will be included. Patients with the diagnosis of head injury or a pathology of the venous sinus without any associated fracture will be excluded. Also, studies whose data cannot be disaggregated to one particular country will be excluded.

#### Intervention

Studies reporting on the diagnosis, and/or treatment (conservative or surgical treatment) for fractures over the venous sinuses will be included. Patients diagnosed with a venous sinus fracture whose method of diagnosis or treatment is unclear will be excluded.

#### Comparison

We shall compare different modalities of treatment across various age groups (adults vs paediatric patients), and income levels according to the World Bank Country and Lending group.^13^

#### Outcome

Studies reporting on (1) mortality rate (2) complication rate (3) Glasglow Outcome Score (4) length of hospital admission (5) symptoms resolved/reduced/ unchanged/worsened will be included.

#### Types of studies

Will be included peer-reviewed publications such as: Cross-sectional studies, case reports, case series, cohort studies, trials and audits. There will be no language restriction and where needed, reports will be translated using online translation platforms such as DeepL Translator (https://www.deepl.com/en/translator). The following articles will be excluded if they do not have disaggregated data on fractures over the venous sinus. Perspectives, letters, opinion pieces, editorials, comments, guidelines, reviews, meta-analyses and articles published in non-peer-reviewed journals will be excluded.

### Stage 3: Study selection

The search results will be uploaded to Rayyan (https://www.rayyan.ai) to facilitate de-duplication and independent, blinded screening.^14^ Initially, titles and abstracts will be screened by two independent reviewers against the eligibility criteria. This will be followed by full-text screening where the same reviewers will screen full-text studies using the eligibility criteria mentioned above. Prior to these two steps, a calibration exercise will be carried by the screeners to ensure an adequate understanding of the eligibility criteria. During the full-text screening phase, corresponding authors of articles will be contacted if the full-text cannot be accessed. A reminder will be sent after 2 weeks If no response is received. If still no avail, the article will be excluded and tagged ‘full-text unavailable’. Unless otherwise stated, disagreements will be discussed and resolved amongst the screeners following unblinding of their decisions on Rayyan. Where consensus cannot be achieved, the lead or senior author (BT or IE) will arbitrate. A PRISMA flow diagram (**figure 1**) will be presented reflecting the search process.

### Stage 4: Charting the data

Data from studies who met the inclusion criteria will be independently extracted and checked by two members of the review team using a standardised extraction proforma. Any unresolved disagreements during this stage will be settled by a third reviewer (BT or IE). This stage will be preceded by a pilot phase. In the pilot phase, all extractors will extract data from the same ten randomly selected included articles. This will help ensure the reliability of the data extraction sheet and that all authors accurately, and homogeneously extract the data. Following this phase, necessary changes will be made to the proforma to ensure that it reflects the included studies before initiation of the extraction proper stage. Data items will include information on study and sample characteristics; type, site and aetiology of venous sinus fracture; clinical presentation and method of diagnosis; treatment mode, timing, and follow-up; and primary and secondary outcomes. If data is insufficiently reported, we will contact the corresponding author as described in ‘**Stage 3**’. All extracted data will be recorded and stored in a spreadsheet. The expected data to be extracted are outlined in **Box 2**.

#### Box 2.

Key data to be extracted on the spreadsheet

##### Study characteristic

- First author
- Year of publication
- Country where the study was conducted
- Language of publication
- Year(s) of data collection
- Income level of study setting
- Region of study setting
- Study design
- Number of centers involved
- Number of with fracture over dural venous sinus
- Age (Mean, median, standard deviation and/or range)
- Sex (Female, male)

##### Characteristics of FOVS

- Mechanism of injury
- Type of fracture
- Fracture site
- Associated cranio-cerebral lesions

##### Management of FOVS

- Clinical presentation
- Method of diagnosis
- Treatment mode (Surgical vs non-surgical)
- Timing of treatment

##### Outcome of FOVS

- Mortality rate
- Complication rates
- Glasgow outcome score
- Functional outcome
- Length of hospital admission
- Symptoms resolved/ reduced/ unchanged/ worsened

### Stage 5: collating, summarising and reporting the results

Following extraction, data will be transferred to SPSS V.26 for analysis. Results will be primarily reported in a tabular form showing descriptive statistics. When appropriate, a narrative description of the results will be presented by grouping the data into meaningful summaries to allow comprehensive reporting of the findings. Pooled statistics will be calculated and presented using measures of central tendency and spread. Unless otherwise stated, the statistical significance will be set at p<0.05. The tables and figures may display information such as: The publication year of the studies, the geographical area where the studies were conducted, study design and demographic characteristics of included patients; aetiology, site, and type of venous sinus fracture; clinical presentation, mode of diagnosis, and mode of treatment; the length of hospital stay, timing of follow-up and outcomes.

### Risk of bias assessment

Given the limited and varied evidence on the topic, the proposed scoping review seeks to offer a comprehensive overview of the management and outcomes of venous sinus fractures. As a result of the inherent standard biases associated with new areas of clinical research, a formal bias risk assessment was judged unnecessary.

### Ethics and dissemination

Because this study did not directly involve human individuals, ethical approval was not necessary. Dissemination strategies will include publication in a peer-reviewed journal, oral and poster presentations at local, regional, national and international conferences and promotion over social media. This work will provide an overview of the available evidence on the management and outcomes of venous sinus fractures globally. This will help in the identification of key research priorities and uncovering any disparities among countries. The proposed review may inform the development of new research questions as well as interventions to systematically improve the management and outcomes of patients sustaining a venous sinus fracture.

### Patient and public involvement

None

## ACKNOWLEDGEMENTS

None

## TWITTER

@BerjoDongmo, @PraiseWah, @ngouanfojosiane, @UbraineWunde, @LASebopelo, @ignatiusesene

## AUTHOR CONTRIBUTIONS

BT, and IE were responsible for conceiving the article. IE is the guarantor. BT, UWN, PSW, LS, and JNT wrote the manuscript. IE, BT, UWN, PSW, LS, JNT, and ESB provided a critical appraisal of the manuscript. All authors critically revised and approved the final manuscript.

## REFERENCES

1. Tsai Y-C, Rau C-S, Huang J-F, et al. The association between skull bone fractures and the mortality outcomes of patients with traumatic brain injury. Emerg Med Int. 2022;2022:1296590.

2. UpToDate. https://www.uptodate.com/contents/skull-fractures-in-adults (accessed 17 November 2023)

3. Skull Fracture - an overview | ScienceDirect Topics. https://www.sciencedirect.com/topics/psychology/skull-fracture (accessed 17 November 2023)

4. Wang W, Lin J, Luo F, et al. Early Diagnosis and Management of Cerebral Venous Flow Obstruction Secondary to Transsinus Fracture after Traumatic Brain Injury. J Clin Neurol. 2013;9:259–68.

5. Kim Y-S, Jung S-H, Lim D-H, et al. Traumatic Dural Venous Sinus Injury. Korean J Neurotrauma. 2015;11:118–23.

6. Aziz MM, Molla S, Abdelrahiem HA, et al. Depressed Skull Fractures Overlying Dural Venous Sinuses: Management Modalities and Review of Literature. Turk Neurosurg. 2019;29:856–63.

7. Abdelaal MA, Saro ASEM, Fadl KN, et al. Management of Compound Depressed Fractures Over Major Cranial Venous Sinuses. The Egyptian Journal of Hospital Medicine. 2021;83:1177–82.

8. Qureshi AI, Sahito S, Liaqat J, et al. Traumatic Injury of Major Cerebral Venous Sinuses Associated with Traumatic Brain Injury or Head and Neck Trauma: Analysis of National Trauma Data Bank. J Vasc Interv Neurol. 2020;11:27–33.

9. Mohamed a. Thabit MD. Venous Sinus Thrombosis Following Repair of Compound Depressed Fractures Overlying Sinuses. The Medical Journal of Cairo University. 2019;87:3277–82.

10. Stein SC. The Evolution of Modern Treatment for Depressed Skull Fractures. World Neurosurg. 2019;121:186–92.

11. Arksey H, O’Malley L. Scoping studies: towards a methodological framework. International Journal of Social Research Methodology. 2005;8:19–32.

12. Shamseer L, Moher D, Clarke M, et al. Preferred reporting items for systematic review and meta-analysis protocols (PRISMA-P) 2015: elaboration and explanation. BMJ. 2015;349:g7647.

13. World Bank Country and Lending Groups – World Bank Data Help Desk. https://datahelpdesk.worldbank.org/knowledgebase/articles/906519-world-bank-country-and-lending-groups (accessed 17 November 2023)

14. Ouzzani M, Hammady H, Fedorowicz Z, et al. Rayyan—a web and mobile app for systematic reviews. Syst Rev. 2016;5:210.

