## Supplementary Table 1 for "Management and outcomes of fractures over cranial venous sinuses: A scoping review protocol"

**Search terms for all databases**

**Pubmed**

| # | **Query** |
| --- | --- |
| 1 | (((((skull OR head OR crani*)) AND ((fracture))) AND ((dural OR venous))) AND (Sinus*)) NOT ((books OR editorials OR letters OR meta-analysis OR review)) |

**Global Index Medicus**

| # | **Query** |
| --- | --- |
| 1 | (tw:((Skull OR Head OR Crani*))) AND (tw:((Fracture))) AND (tw:((Dural OR Venous) )) AND (tw:(Sinus*)) |

**African Journal Online**

| # | **Query** |
| --- | --- |
| 1 | (Skull OR head OR Crani*) AND (Fracture) AND (Dural OR Venous) AND Sinus* |

**Scopus**

| # | **Query** |
| --- | --- |
| 1 | ( TITLE-ABS-KEY ( ( skull OR head OR crani* ) ) AND TITLE-ABS-KEY ( ( fracture ) ) AND TITLE-ABS-KEY ( ( dural OR venous ) ) AND TITLE-ABS-KEY ( sinus* ) ) AND ( EXCLUDE ( DOCTYPE , "bk" ) OR EXCLUDE ( DOCTYPE , "no" ) OR EXCLUDE ( DOCTYPE , "cp" ) OR EXCLUDE ( DOCTYPE , "le" ) OR EXCLUDE ( DOCTYPE , "ch" ) OR EXCLUDE ( DOCTYPE , "re" ) OR EXCLUDE ( DOCTYPE , "ed" ) ) AND ( EXCLUDE ( EXACTKEYWORD , "Animal Model" ) OR EXCLUDE ( EXACTKEYWORD , "Histology" ) OR EXCLUDE ( EXACTKEYWORD , "Animal Tissue" ) OR EXCLUDE ( EXACTKEYWORD , "Antibiotic Agent" ) OR EXCLUDE ( EXACTKEYWORD , "Physiology" ) OR EXCLUDE ( EXACTKEYWORD , "Nonhuman" ) ) AND ( LIMIT-TO ( PUBSTAGE , "final" ) ) AND ( EXCLUDE ( SRCTYPE , "k" ) OR EXCLUDE ( SRCTYPE , "Undefined" ) ) |

**Embase**

| **#** | **Query** |
| --- | --- |
| 1 | 'skull'/exp OR skull or 'head'/exp OR head or crani* |
| 2 | 'fracture'/exp OR 'fracture' |
| 3 | venous OR dural |
| 4 | sinus* |
| 5 | 1 AND 2 AND 3 AND 4 |
| 6 | limit 5 to humans |
| 7 | limit 6 to articles |
